# Booster protection against Omicron infection in a highly vaccinated cohort

**DOI:** 10.1101/2022.02.24.22271347

**Authors:** Caroline G. Tai, Lisa L. Maragakis, Sarah Connolly, John DiFiori, Leroy Sims, Eleanor Adams, Deverick J. Anderson, Michael H. Merson, David D. Ho, Yonatan Grad, Christina DeFilippo Mack

## Abstract

**Background:** Evaluation of COVID-19 vaccine booster effectiveness is essential as new variants of SARS-CoV-2 emerge. Data support the effectiveness of boosters in preventing severe disease and hospitalization; however, the real-world impact on reducing incident SARS-CoV-2 infections across specific variants is not yet clear.

**Objectives:** Assess the impact of COVID-19 boosters on protection against SARS-CoV-2 infection in a real-world setting from a highly vaccinated (99%) population curated from a linked database with test results, vaccination history, patient demographics, and genomic sequencing information from the National Basketball Association (NBA).

**Methods:** In this population, 1,613 fully vaccinated staff and players (median age 34.5 years, 88% male) who tested at least once between 1 December 2021 and 5 January 2022, were analyzed. Individuals were tested at the time of reporting any symptom, regardless of severity, after known exposure per self-report or contact tracing and/or during enhanced surveillance. Boosted individuals (n=1260) were compared to fully vaccinated and booster eligible individuals (n=162), defined as 2 months post-JNJ-78436735 or 5-months post-second dose of mRNA vaccine. Individuals not yet eligible for a booster and those who recovered from COVID-19 between 1 November and 30 November, 2021 (n=25) were examined in secondary analyses but excluded from the primary comparison. Individuals who were not fully vaccinated (n=27) or who received a booster within 14 days during or prior to the observation period (n=916) were excluded.

**Results:** In this closely monitored population, fully vaccinated booster-eligible individuals were 2.6 times more likely (RR = 2.6, 95% CI: 2.2 to 3.0, p<0.0001) to have a confirmed COVID-19 infection than boosted individuals. Secondary analysis including non-boosted individuals with recent vaccination or recent SARS-CoV-2 infection found that non-boosted individuals were at greater risk of infection compared with boosted individuals (RR = 2.1, 95% CI: 1.8 to 2.4, p<0.001). Results were similar when stratified by primary vaccination type with overlapping confidence intervals. No hospitalizations or deaths were observed in this cohort. Genetic sequencing confirmed 93% of infections to be Omicron (among n=330 sequenced).

**Conclusions:** These results highlight the protective benefit of boosters against incident SARS-CoV-19 infection, not only for severe symptoms and death. Assessment of booster effectiveness remains vital for COVID-19 vaccines as new variants of SARS-CoV-2 emerge, as there is still uncertainty around performance in real-world settings. These data are needed to convince the general public that boosters remain effective at preventing in the spread of COVID-19.

## Manuscript

Evaluation of COVID-19 vaccine booster effectiveness is essential as new variants of SARS-CoV-2 emerge. Data support the effectiveness of boosters in preventing severe disease and hospitalization; however, the impact on reducing incident SARS-CoV-2 infections is not yet clear [1-3]. Data from the National Basketball Association (NBA) assessed the impact of COVID-19 boosters on protection against SARS-CoV-2 infection in a highly vaccinated (99%) population.

In this population, 1613 fully vaccinated (as defined by CDC guidelines[4]) staff and players (median age 34.5 years, 88% male) who tested at least once between Dec 1, 2021 and Jan 5, 2022, were analyzed (Table 1). Individuals were tested at the time of reporting any symptom, regardless of severity, after known exposure per self-report or contact tracing and/or during enhanced surveillance. Boosted individuals (n=1260) were compared to fully vaccinated and booster eligible individuals (n=162), defined as 2 months post-JNJ-78436735 or 5-months post-second dose of mRNA vaccine [4]. Individuals not yet eligible for a booster and those who recovered from COVID-19 between Nov 1 and Nov 30, 2021 (n=25) were examined in secondary analyses but excluded from the primary comparison. Individuals who were not fully vaccinated (n=27) or who received a booster within 14 days during or prior to the observation period (n=916) were excluded.

**Table 1:** Booster protection against SARS-CoV-2 infection, Dec 1, 2021 - Jan 5, 2022.

| Population Characteristics | Overall | Boosted | Non-Boosted |
| --- | --- | --- | --- |
| Total number of fully vaccinated individuals <sup>a</sup> | 1613 | 1267 | 346<br>(180 booster eligible) |
| Male, % | 88% | 87% | 93% |
| Median age, years (IQR) | 34.5 (27.8 - 46.4) | 37.6 (30.0 - 48.7) | 27.1 (23.6 - 32.3) |
| Median number of tests per individual (IQR) | 14.0 (8.0 - 22.0) | 14.0 (8.0 - 22.0) | 16.0 (9.0 - 23.0) |
| Individuals with recent infections during Nov 1, 2021 – Nov 30, 2021, # (%) | 25 (2%) | 7 (1%) | 18 (5%) |
| Individuals with COVID-19<sup>b</sup> during observation period Dec 1, 2021 – Jan 5, 2022, # (%) | 479 (30%) | 304 (24%) | 175 (51%) |
| Primary Vaccination Type, # (%) |  |  |  |
| mRNA | 1262 (78%) | 965 (76%) | 297 (86%) |
| Viral vector | 351 (22%) | 302 (24%) | 49 (14%) |
| **Comparisons:** |  | **Risk Ratio (95% CI)** | **P-value** |
| **Non-boosted, but booster eligible (n=162) versus boosted (n=1260), excluding recent infections in Nov 2021** |  | **2.6 (2.2, 3.0)** | **p<0.0001** |
| Stratified by primary vaccination type: |  |  |  |
| mRNA |  | 2.8 (2.3, 3.3) |  |
| Viral Vector |  | 2.3 (1.8, 2.9) |  |
| **Non-boosted (n=346) versus boosted (n=1267)** |  | **2.1 (1.8, 2.4)** | **p<0.0001** |
| Stratified by primary vaccination type: |  |  |  |
| mRNA |  | 2.26 (1.92, 2.67) |  |
| Viral Vector |  | 2.01 (1.53, 2.64) |  |

<sup>a</sup> Fully vaccinated was defined as two doses of an mRNA vaccine (BNT162b2 or mRNA-1273) or 1 dose of JNJ-78436735 per CDC guidelines [4].
<sup>b</sup> Diagnostic testing for COVID-19 was conducted via a molecular test using mid-turbinate swab with the Cue Health system or Mesa Accula test platform or pooled nasal oropharyngeal swab with the Roche cobas platform to detect the presence RNA of SARS-CoV-2. All positive tests were confirmed with a subsequent test on a Roche cobas platform.

In this closely monitored population[5], fully vaccinated booster-eligible individuals were 2.6 times more likely (RR = 2.6, 95% CI: 2.2 to 3.0, p<0.0001) to have a confirmed COVID-19 infection than boosted individuals. Secondary analysis including non-boosted individuals with recent vaccination or recent SARS-CoV-2 infection found that non-boosted individuals were at greater risk of infection compared with boosted individuals (RR = 2.1, 95% CI: 1.8 to 2.4, p<0.001). Results were similar when stratified by primary vaccination type with overlapping confidence intervals (Table 1). No hospitalizations or deaths were observed in this cohort. Genetic sequencing confirmed the Omicron variant to be the dominant, nearly exclusive variant, in this sample period, with 93% confirmed to be Omicron (among n=339 with genomic sequencing).

Overall, this study is a younger, healthier cohort compared to the U.S. population, precluding generalizability. There was more frequent testing among the non-boosted group (Table 1, median number of tests per person) and as daily surveillance testing was not performed, it is possible that some asymptomatic cases or unreported symptomatic cases were not detected. These results highlight the protective benefit of boosters against SARS-CoV-19 infection, specifically reducing incident infection with the Omicron variant.

## Data Availability

Data produced in the present study are available upon reasonable request to the authors if approved by research committee governing these protected occupational health data records.

